# Developing an AI-Enhanced Individualized Prediction Tool for Psychopathological Symptoms in Vietnam: A Study Protocol

**DOI:** 10.1101/2025.08.03.25332928

**Authors:** Hung Nguyen, Minh Khau, Huyen Nguyen, Hieu Pham

## Abstract

Artificial intelligence (AI) is increasingly leveraged in mental healthcare for early detection, monitoring, and personalized intervention. However, most existing AI applications are based on categorical diagnostic systems like DSM-5 or ICD-11, which often lead to comorbidity issues, ambiguous diagnoses, and insufficient personalization. These tools typically target specific disorders (e.g., depression or anxiety), neglecting the broader, interconnected nature of psychopathological symptoms. Addressing these limitations, recent innovations in psychopathology emphasize transdiagnostic and network-based approaches, such as the Hierarchical Taxonomy of Psychopathology (HiTOP), which conceptualize mental disorders as dimensional and inter-connected constructs. This study proposes an AI-powered tool that integrates data-driven principles from both the HiTOP and symptom network models to generate individualized risk profiles for internalizing mental disorders (e.g., depression, anxiety, bipolar disorders). Our solution aims to assess individuals’ current psychopathological traits and symptom components, providing a comprehensive, nuanced profile supporting clinical diagnoses and monitoring.

The study unfolds in three phases: (1) model ideation; (2) model implementation in a large-scale Vietnamese sample; and (3) deployment in clinical and psychological practice settings in Vietnam. Central to our method is the development of a Risk-aware Taxonomy-enhanced Symptom Encoder (RiTaSE), which encodes symptom data and their severities into rich representations processed via a Transformer-based model. The model is trained using high-quality, validated datasets mapped to the HiTOP framework.

This project is among the first to employ AI for personalized psychopathological profiling in Vietnam,, as well as other low- and middle-income countries. Expected outcomes include an advanced diagnostic-support tool for clinical use, improved crosscultural insights into symptom comorbidity, and practical utility in mental health monitoring and intervention evaluation. Future extensions aim to broaden the scope across all HiTOP dimensions and predict transitions to clinical states through longitudinal and multi-modal data integration.

## I. Introductions

Mental health issues are increasingly recognized as a critical public health concern in Vietnam, with increasing prevalence rates of depression, anxiety, and related disorders post Covid-19, particularly among adolescents and young adults [1], [2]. However, access to mental health care remains severely limited due to a shortage of trained professionals, stigma surrounding mental illness, and uneven distribution of services across urban and rural areas [3], [4]. The conventional model of care, relying heavily on face-to-face sessions with mental health professionals, is insufficient to meet the growing demand for timely diagnosis, monitoring, and intervention [5]. This gap places increased burden on mental health practitioners in Vietnam, contributing to high rates misdiagnosis, underdiagnosis, and relapse, while also posing challenges in tracking remission and long-term recovery.

In response to these challenges, digital and artificial intelligence (AI)-driven solutions are gaining attention internationally for their potential to detect, monitor, and personalize mental health interventions [6]. In Vietnam, however, the integration of AI into mental healthcare remains in its infancy with recent studies implementing machine learning in detecting risks of increased stress [7] or depression [8]. Nonetheless, these studies are limited in cultural adaptation of psychological measurements and integration of advancements in psychopathological theories. Moreover, most AI applications in both Vietnam and globally target specific mental disorders such as depression and anxiety [3], limiting their capacity to handle comorbidity, individual variability, and transdiagnostic patterns of psychopathology.

To address these limitations and advance mental healthcare in Vietnam, this study proposes a novel AI-powered tool supported by recent advancements in psychopathological theories. Specifically, our tool seeks to generate individualized mental health profiles based on current psychopathological traits and symptom components, offering a comprehensive and detailed approach to inform clinical assessment and ongoing monitoring.

## II. Conceptualization of Psychopathology

Psychopathology is commonly classified using traditional standardized frameworks such as the Diagnostic and Statistical Manual of Mental Disorders (DSM-5) [9] and the International Classification of Diseases, 11th Revision (ICD-11) [10]. While these systems provide a shared language for diagnosis and research, they are often criticized for their categorical structure and rigid boundaries [11]. Specifically, the dichotomous nature of these frameworks, defined mental disorders as either present or absent based on certain criteria and thresholds, overlooks the high comorbidity between disorders and the clinical relevance of subthreshold symptoms [12]. Moreover, the clinical thresholds of disorders are often arbitrary and unstable [13], [14], leading to reliability problems, meaning that different clinicians might have different views on whether a person has a disorder or not. Additionally, overlap of major symptoms can blur distinctions between disorders, leading to inconsistent diagnoses and potential misclassifications [15], [16]. Bipolar disorders, in particular, are frequently misdiagnosed as either depression, anxiety or schizophrenia due to shared symptoms of elevated anhedonia, restlessness and psychosis respectively [17]. In contrast, the DSM-5 and ICD-11 allowing high symptom variability within disorders leads to different symptom profiles among individuals with the same diagnosis, or heterogeneity problems [18], [19]. This complicates treatment planning and contributes to inconsistent responses to intervention, increasing the burden on clinicians.

Furthermore, this traditional framework does not align with scientific observations in the field of neuroscience, biological psychiatry, molecular genetics, and clinical psychology [20], [21]. Particularly, genetics studies on schizophrenia observed less severe conditions among biological relatives of people diagnosed with the disorder [22] while neuro-imaging studies found increased grey-matter loss as schizophrenia progressed [23], suggesting that schizophrenia symptoms exist on a continuum with certain vulnerable genes [24]. Likewise, certain neural circuits are identified to facilitate symptoms of both depression and anxiety [25], [26], explaining the high comorbidity of the two disorders. This mismatch in theoretical approach and scientific findings hinders clinical utility and effective care for people with mental disorders. Therefore, in designing this project, we adopt emerging datadriven frameworks that reflect contemporary advances in the field, mitigating the challenges of traditional approaches.

### A. The Hierarchical Taxonomy of Psychopathology

The Hierarchical Taxonomy of Psychopathology (HiTOP) is a transdiagnostic dimensional approach to psychopathology, viewing mental disorders and symptoms as continuous dimensions instead of a dichotomous perspective. The framework was developed based on empirical patterns of correlated psychopathological phenomena, utilizing quantitative methods inferring latent variables, such as factor analysis and latent class analysis [11], to integrate parallel literature of dimensional maladaptive personality and mental disorders into a unified, overarching hierarchical model [12], [27]. Figure 1, derived from [27], illustrates the HiTOP model, ranging from broader psychopathological dimensions, called spectra, at the upper levels to more specific symptoms, signs and traits at the lower levels.

**Fig. 1.**
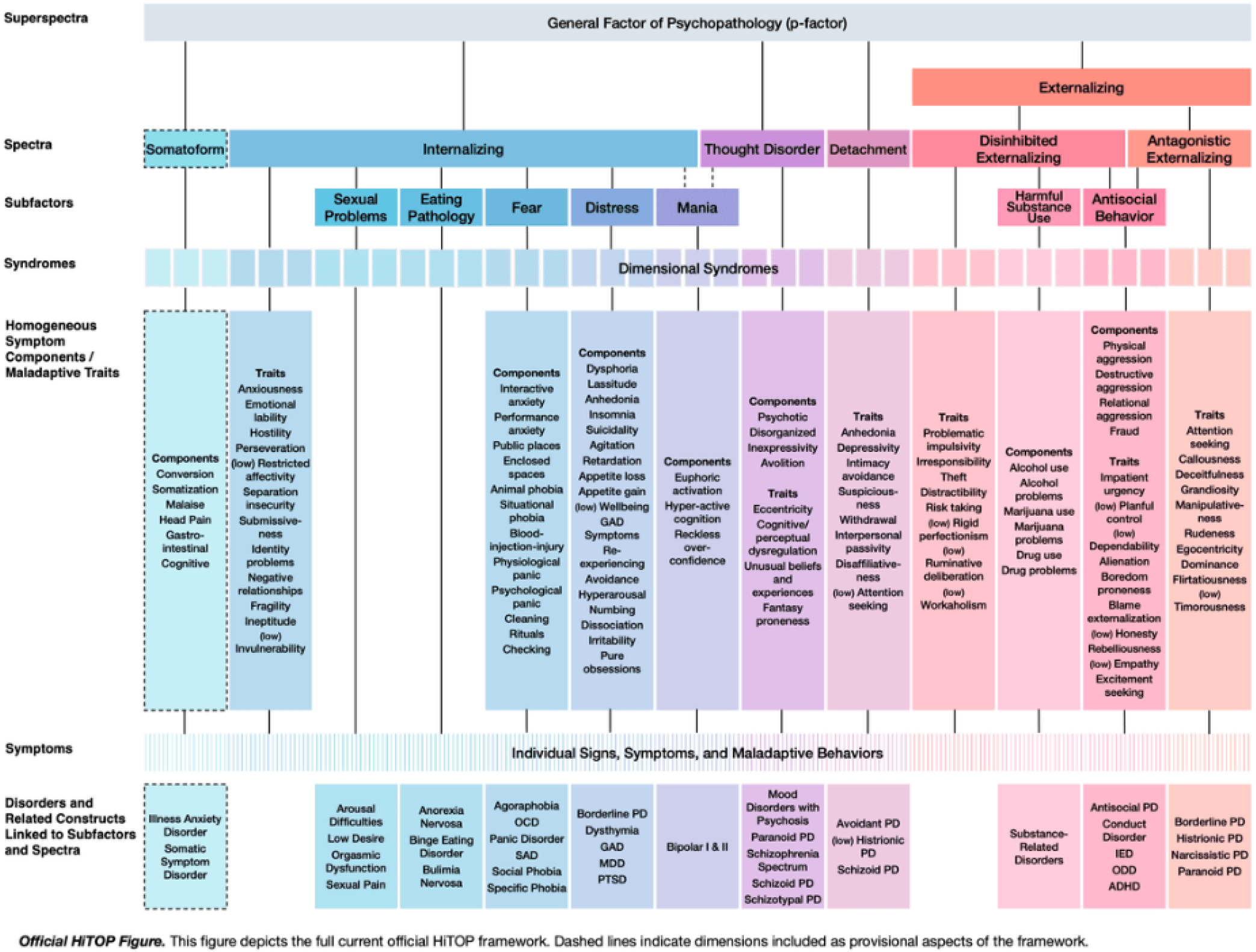
The current HiTOP framework, derived from [27]

Approaching psychopathology from this view, several problems existing in traditional psychopathological frameworks are overcome. Firstly, the dimensional nature of HiTOP better reflects real-world observations that psychopathology generally exhibits no distinct boundaries across varying severity levels [28]. Secondly, HiTOP embraces comorbidity among mental disorders as the framework is developed based on covariation among mental health variables, allowing overlapping among symptoms [11]. Lastly, HiTOP diminishes the dependence on arbitrary diagnostic thresholds due to its use of dimensionality, eliminating issues with subthreshold symptoms [29]. Hence, HiTOP allows diverse psychopathological profiles with minimal restriction to classifications.

### B. The Network Psychometric Approach

Network psychometric is an emerging perspective on psychopathology stemming from the network theory, viewing all psychological phenomena as complex and dynamic biopsycho-social systems [12]. Similarly to the HiTOP framework, network psychometric is a data-driven approach to psychopathology grounded in covariation and co-occurence of symptoms. However, the network approach is underpinned by the application of network theory and statistical network models to psychology instead of HiTOP’s latent variable approach [30]. This approach proposes that psychopathology, or mental disorders, arises from systemic and interconnected interactions between symptoms (Figure 2). Thus, psychopathology progresses over time from a within-person system.

**Fig. 2.**
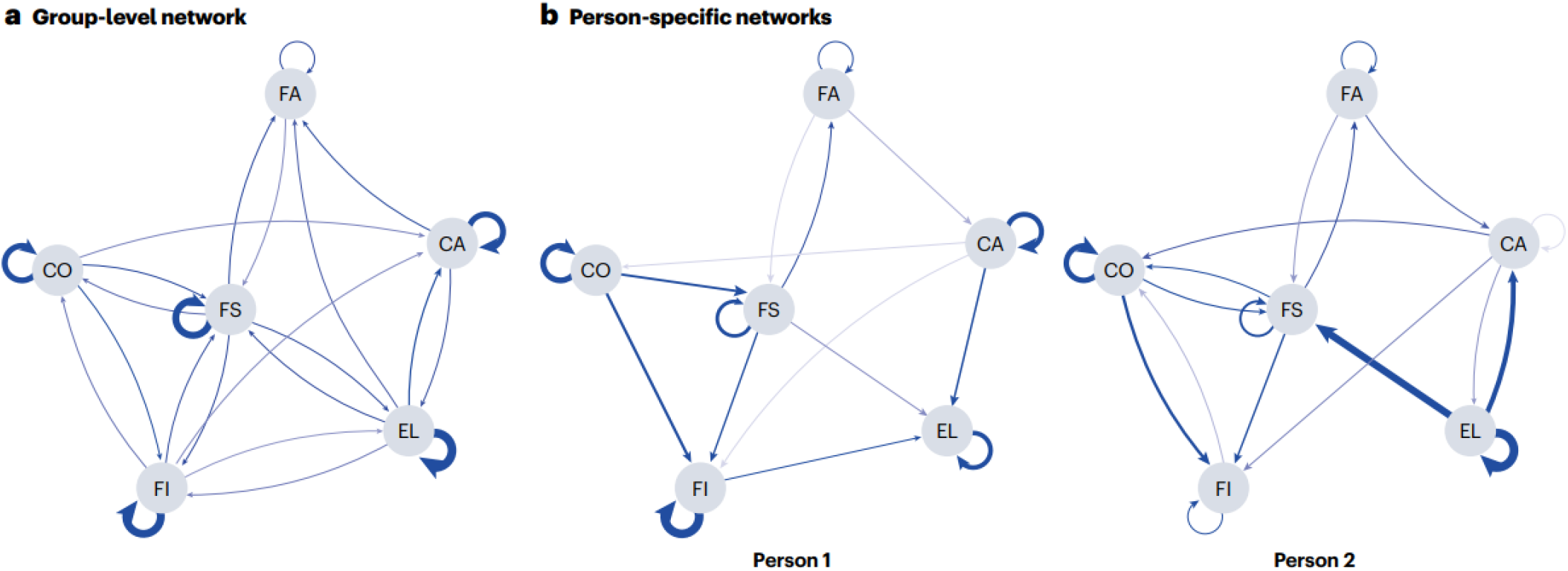
Illustration of the network approach to psychopathology, derived from [12]

Addressing challenges of the traditional framework, the network psychometric offers an alternative perspective on psychopathology. This perspective emphasizes the analysis of symptoms at the individual level and prioritizes their dynamic interactions, thereby shifting the focus from meeting predefined diagnostic criteria to identifying central symptoms within a person’s unique mental health system [31]. This mitigates heterogeneity problems and promotes individualized care, in line with the principles of precision medicine. Furthermore, the network approach highlights that comorbidity, regardless of diagnostic boundaries and thresholds, arises naturally from the causal interactions between symptoms [32], aligning with the empirical evidence of high comorbidity rates among multiple disorders.

### C. Combining HiTOP and Network Psychometric

HiTOP and network psychometric, despite being developed on different statistical and theoretical foundations, are complimentary perspectives on psychopathology by addressing each other’s limitations [12]. Stemming from latent variables perspective, a major assumption of HiTOP is local independence, meaning that the framework overlooks interactions of symptoms from different underlying components and spectra while network psychometrics lacks an psychopathological structure, limiting replication and generalization [33]. Hence, combining the two approaches helps gain a more comprehensive view of individualized psychopathological profiles.

Recent advancements in psychometric modeling have demonstrated the potential of integrating latent variable models with network approaches to gain exploratory insights into the relationships between latent constructs and observable symptoms [34], [35]. This integration offers a more nuanced and comprehensive framework for understanding the structure of psychopathology [33]. One such approach, Exploratory Graph Analysis (EGA), emerges from the network perspective and is designed to identify latent dimensions by detecting clusters of symptoms with high co-occurrence in a network structure [36], [37]. In parallel, methodological developments have introduced frameworks for modeling networks among latent variables or among residual item-level associations after accounting for latent constructs [33]. These innovations contribute to a unified psychometric approach that bridges the gap between traditional latent variable theory and contemporary network analysis.

This study, leveraging the use of artificial intelligence (AI) techniques, attempts to combine network psychometrics and latent variable modeling, particularly the HiTOP framework, to develop a tool for predicting individualized psychopathological profiles. Specifically, we propose an AI model that estimates a broad range of symptom severities from the limited observed symptom data by identifying interaction patterns and network structures within the internalizing spectrum of HiTOP, focusing on a Vietnamese sample.

## III. The Individualized Risk Profile Prediction Model for Psychopathology Symptoms

### A. Overview

#### 1) Problem Formulation

Over a set of symptoms Ψ, given a psychopathology profile *X* = {*µ*_*u*_ | *u* ∈ Ψ}, where *µ*_*u*_ ∈ [0, 1] is the score, or severity level, or risk level, of symptom *u*, refine *X* into 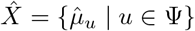where 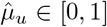 is the risk refined by the model of symptom *u*, based on the latent comorbidity pattern between them.

Based on this formulation, the input profile *X* is from the raw data such as self-report questionnaires. In real-world datasets, the symptom risk level is often missing as a single self-report questionnaire could not address all symptoms but a subset of which are being focused on. Therefore, a missing-risk symptom will be marked with a binary indicator (is missing or not) and the risk level is set to 0. Then, the model is tasked with refining raw risk levels and generating risk levels for the missing.

The HiTOP taxonomy *T* ={ *V, E*} where *V* ⊇ Ψ is the set of psychopathology concepts in the HiTOP framework and *E* ={⟨; *u, v*⟩ | *u, v* ∈ *V*} is the set of direct hierarchies between the concepts *u* and *v*. Note that in the taxonomy, the set of concepts *V* may contain ones that are not in the symptom set Ψ. In this problem, the items in the set Ψ are at the same level of hierarchy in the HiTOP taxonomy *T*, which specifically is the symptom level.

#### 2) Intuition

The network approach of psychopathology hypothesizes a directed partial correlation ⟨*u, v*⟩ in which a symptom *u* interacts with a symptom *v* and contributes to *v*’s activation. These interactions are latent and need to be learned. In the meantime, the HiTOP framework includes relationships between psychopathology concepts, forming a tree-like graph. In this graph, a symptom *u* (e.g. insomnia) that can be reached from a latent variable *c* (e.g. distress subfactor) is said to have empirically higher co-occurrence with a symptom *v* (e.g. chronic fatigue) that can also be reached from *c* than any other symptom *w* that cannot be reached from *c* (e.g. substance abuse - as under the disinhibited externalizing subfactor). These observations suggest that we can harness the structure of the HiTOP framework in addition to the collected dataset to learn about the latent interactions between the symptoms.

#### 3) Model Architecture

Building on this intuition, we propose a model called **Ri**sk-aware **Ta**xonomy-enhanced **S**ymptom **E**ncoder (RiTaSE, Figure 3), a novel framework designed to generate contextual symptom embeddings by integrating dynamic risk states with structural insights from the HiTOP taxonomy. RiTaSE achieves this through two synergistic components: 1) The taxonomy representation learning module that constructs a position-aware ego network to encode hierarchical and semantic relationships within the HiTOP framework, preserving both structural topology and node-specific attributes, 2) The symptom comorbidity learning module leveraging a Transformer-based encoder [38] that synthesizes risk-contextualized interactions between symptoms, producing embeddings that reflect both individual risk profiles and systemic comorbidity patterns.

**Fig. 3.**
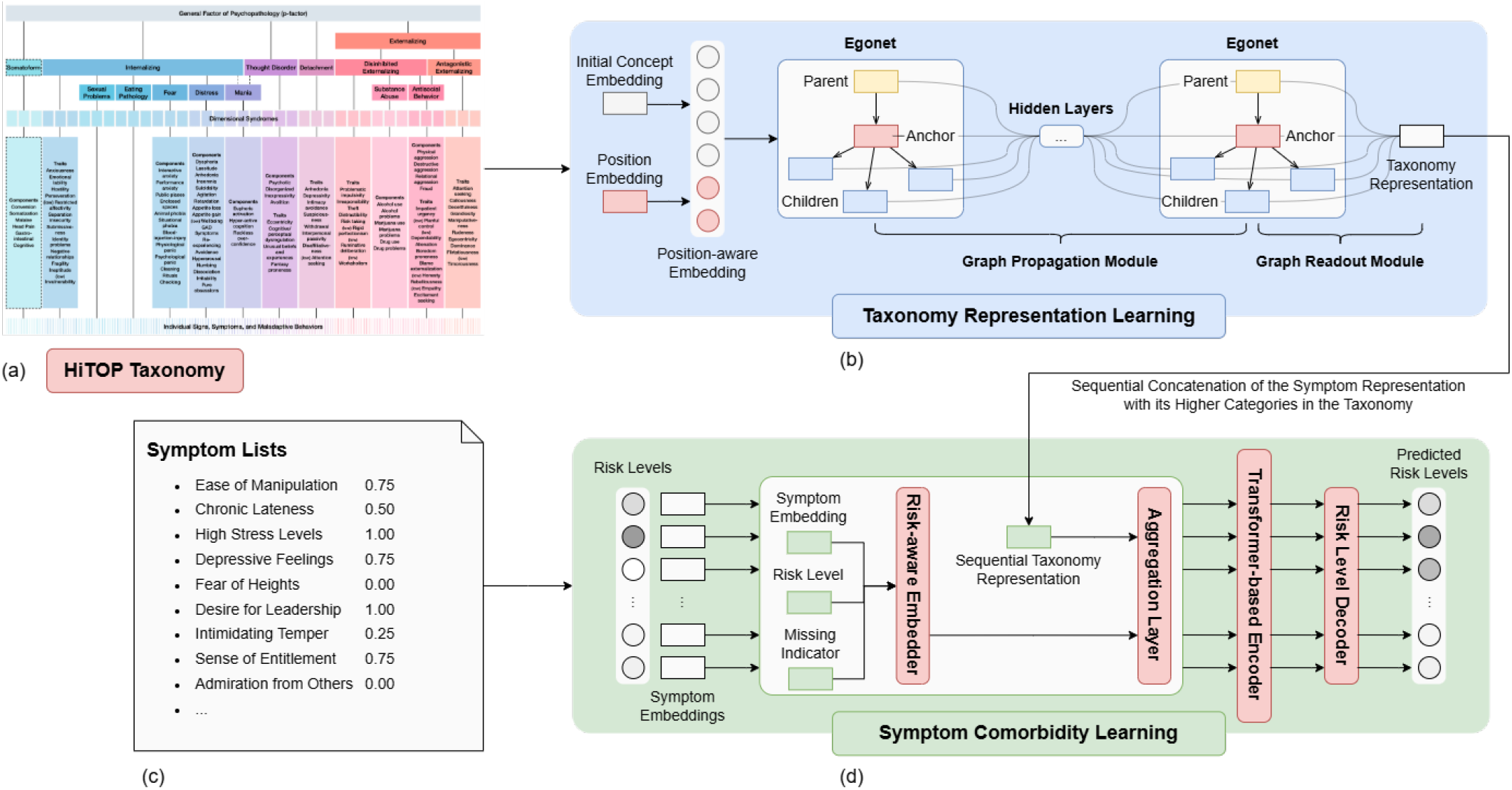
Overview of the model architecture. a) The HiTOP taxonomy derived from [27]. b) The taxonomy representation learning module. c) The input psychopathology profile *X*. d) The symptom comorbidity learning module.

### B. Taxonomy Representation Learning

The taxonomy representation module employs ego-network construction for each HiTOP concept, following methodologies established in TaxoExpan [39]. For each concept *c*, we generate a position-aware ego-network where node embeddings are augmented with structural positional encodings. These enriched representations undergo propagation through a graph neural network (GNN) to capture multi-hop hierarchical dependencies. A readout operator subsequently aggregates neighborhood features into a single representation for the central concept of the ego-network, preserving both structural semantics and node attributes.

### C. Symptom Comorbidity Learning

This component integrates risk-aware symptom features with taxonomic context through a multi-stage process:

- First, symptom embeddings are modulated by their current risk profiles to produce risk-aware symptom embeddings.
- Then, these embeddings are aggregated with their corresponding HiTOP taxonomy representations, creating fused vectors encoding symptom risk, taxonomic position, and intrinsic symptom properties and comorbidity pattern.
- A Transformer-based encoder employs a multi-head selfattention mechanism to process the fused embeddings, generating contextual symptom representations through cross-symptom attention patterns.
- With all of that synthetic information embedded, a decoder is trained to map final representations to refined risk level estimates.

### D. Learning and Inference

The taxonomy representation learning module and the symptom comorbidity learning module are trained separately. First, the taxonomy representations are trained in a self-supervised discipline. Specifically, for each concept *c*_*q*_ in the taxonomy, we construct a contrastive dataset *D*_contrast_ with its parent node *c*_*p*_ as positive example and *M* randomlyselected other nodes 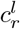 which are neither the parents nor the descendants of *c*_*q*_, as negative examples. Those nodes serve as anchor nodes while *c*_*q*_ is the query node.

Given the constructed self-supervised dataset 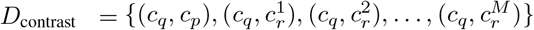 the model need to distinguish the valid tuple from the rest. This is done using an InfoNCE loss [40]:

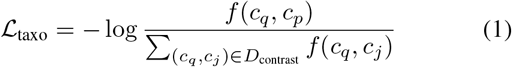

where *f* (·) is the matching module, functioning as a log-bilinear model, *j* = 1, 2, …, *M* + 1, and ℒ_taxo_ is the cross entropy of classifying the positive pair (*c*_*q*_, *c*_*p*_) correctly.

For the symptom comorbidity learning module, inspired by the masked language modeling method [41] that successfully model word co-occurrence statistics, we attempt to train the module by stochastically masking *p*_mask_% of symptom risks in each profile. Specifically, the masked symptom’s risk level is set to 0 and the missing indicator is set to 1 (missing); or the masked symptom is set to be in a “perturbed” setting in which the missing indicator is kept as 0 (not missing) but the risk level is added to/subtracted from a perturbation value. The decision will based on a distribution that we will determine later. Then, the model need to reconstruct the masked values via mean squared error minimization:

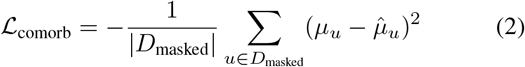

where *D*_masked_ is the set of masked symptoms.

### E. Evaluation Metrics

To assess model performance rigorously, we employ three metrics that quantify prediction accuracy and explanatory power. Experimental implementation details are provided in Section IV.

#### 1) Mean Absolute Error

The Mean Absolute Error (MAE) metric measures the average prediction accuracy by calculating the absolute difference between predicted symptom risk levels 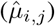 and actual observed values (*µ*_*i,j*_) across all profiles and symptoms.

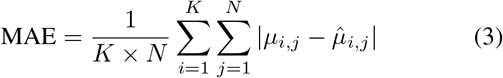

where *K* represents the number of symptoms and *N* represents the total number of profiles in the evaluation set. Lower MAE values indicate better model performance, with MAE approaching zero representing perfect prediction accuracy.

#### 2) Averaged Median Absolute Error

While MAE offers clear interpretability, it can be affected by extreme values in the data. To address this, we also use the averaged Median Absolute Error (avgMedAE), which calculates the median of the absolute differences between predicted and actual values across all profiles for each symptom and then averages them. This approach reduces the impact of outliers, providing a more robust assessment of model performance.

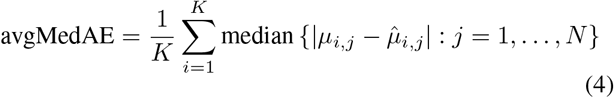

#### 3) Averaged R-squared

This metric quantifies the model’s explanatory power by measuring how well it captures variance in symptom risk levels. Considering each symptom’s risk level as an individual prediction target, we compute a separate R^2^ value for each symptom and then average these values to obtain an overall performance measure:

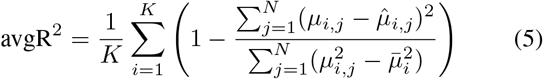

where 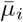 represents the mean risk level for symptom *i* across all evaluation profiles. This averaged R^2^ (avgR^2^) metric indicates the proportion of variance in symptom risk levels that the model successfully explains using other symptoms as contextual predictors. Higher avgR^2^ values (closer to 1.0) indicate that the model effectively leverages symptom interdependencies to predict individual symptom risk levels.

## IV. Project Design

The research project is structured into three distinct phases, as depicted in Figure 4:

**Fig. 4.**
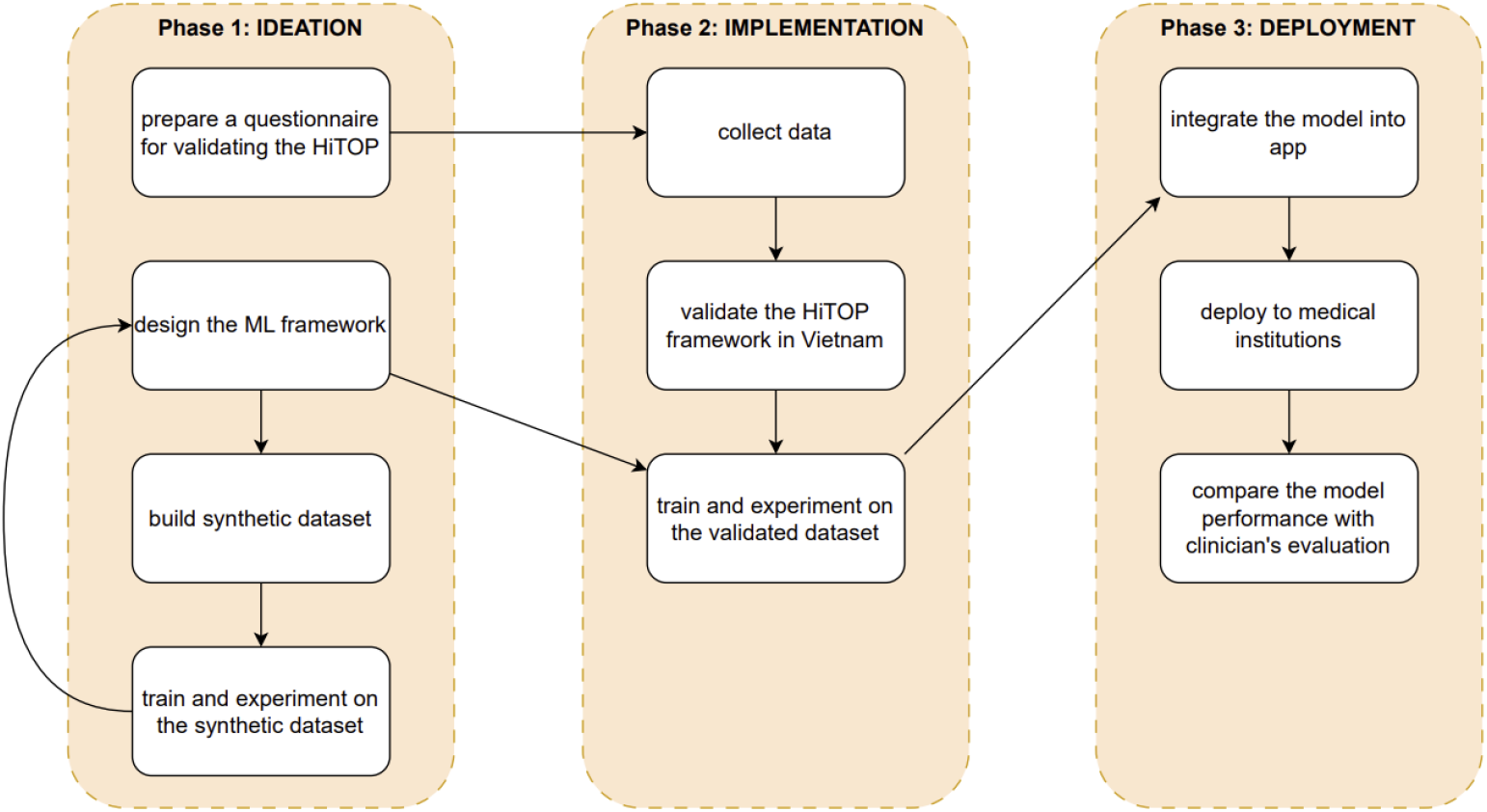
Project pipeline with specific action items. Arrows indicate flows of items to execute in the project.

- **Phase 1: Ideation** focuses on developing the ML model. This involves designing the ML framework, creating a synthetic dataset, conducting initial experiments using this dataset, and preparing a questionnaire for validating the HiTOP system in the next phase.
- **Phase 2: Implementation** centers on data collection and the validation of theoretical constructs. In this phase, data is gathered and the HiTOP framework is validated within the Vietnamese context. The validated dataset is then used to train and validate with the model.
- **Phase 3: Deployment** addresses the integration and application of the AI tool. The trained model is incorporated into an application, deployed to medical institutions, and its performance is compared with clinician evaluations to assess diagnostic process performance both with and without the assistance of the proposed tool.

### A. Phase 1 - Ideation

The ideation phase serves as the foundational stage for developing an AI-enhanced individualized prediction tool for psychopathology assessment using the HiTOP framework. This phase encompasses the following interconnected components that work synergistically to establish the technical and methodological groundwork for subsequent phases:

#### 1) Model Design

The ML model development follows an iterative approach that allows for continuous refinement of the architecture proposed in Section III. During this process, the core algorithmic components are designed and tested, with particular emphasis on the model’s ability to learn complex comorbidity patterns and predict unmeasured symptom dimensions.

#### 2) Synthetic Dataset Construction

The synthetic dataset construction utilizes predefined comorbidity patterns or draws from published datasets that implement the HiTOP framework. The synthetic data generation process integrates well-documented psychopathological relationships and comorbidity associations, establishing a controlled computational environment that supports rapid prototyping and iterative testing. This approach allows for thorough model testing and improvement without needing real patient data at the early development stage.

#### 3) Questionnaire Development for Dataset Validation

Concurrently, a comprehensive questionnaire is being developed to validate the HiTOP framework within the Vietnamese population. The HiTOP internalizing spectrum, preliminarily developed and finalized by Watson et al. [42], is selected due to its wide coverage across multiple common mental disorders and our limited capacity. The measurement contains 39 subscales with 213 items evaluating a wide range of internalizing psychopathological traits such as anxious worry, anhedonic depression and traumatic reaction. Responses are gathered based on a 4-point Likert scale, rating items from 0 to 3 with 0 being *“not at all”* and 3 being *“a lot”*. Items are statements indicating the presence or absence of mental health problems. *“I was overwhelmed by anxiety”* is an example of an item in anxious worry subscale. The measurement is forward and backward translated by at least 2 different bilingual translators as suggested by Cruchinho et al. [43].

### B. Phase 2 - Implementation

This phase aims to train and experiment our AI model on a Vietnamese dataset, utilizing a validated HiTOP’s internalizing spectrum for Vietnamese populations. Thus, in this phase, we aim to gather a large-scale and robust dataset of Vietnamese participants for framework validation and model training.

#### 1) Procedure

A total of 1,000 participants aged 18 and above will be recruited, with approximately 20% to 30% from clinical settings and the remaining from the general population, reflecting estimated rates of mental health disorders in the broader population of Vietnam [2], [44]. Clinical participants will be recruited from psychiatric hospitals and outpatient clinics by collaborating with trained practitioners. Non-clinical participants will be recruited online via social media, university mailing lists, and survey platforms.

After providing informed consent, participants will complete demographic questions and the HiTOP-SR internalizing module. Clinical participants will complete paper-based questionnaires; non-clinical participants will complete them online. Data quality will be ensured through embedded attention checks and response time thresholds. Ethical approval will be obtained prior to data collection.

#### 2) Measurement

We collect participants’ demographic information, including age, sex, sexual orientation, geographic and cultural background, education levels, socioeconomic status, income level, and history of mental health. Regarding psychometric measurement, the HiTOP internalizing questionnaire, described in Phase 1, will be used to assess psychopathological traits.

#### 3) Statistical Analysis

Statistical analysis for HiTOP validation and adaptation is performed in R version 4.3.3 and partly in IBM SPSS 26. Model training and experiment on validated Vietnamese data and HiTOP framework are carried out in Python using PyTorch Lightning [45] for code organization and model training and Weights & Biases [46] for experiment tracking and visualization.

Demographic characteristics will be summarized using descriptive statistics. In line with previous research adapting the HiTOP framework to the German population [47], unidimensional congeneric confirmatory factor analyses (CFAs) will be conducted to evaluate the internal structure of each scale. Model fit will be assessed using multiple indices, including the unbiased Standardized Root Mean Squared Residual (uSRMR), Comparative Fit Index (CFI), Root Mean Squared Error of Approximation (RMSEA), Akaike Information Criterion (AIC), and Bayesian Information Criterion (BIC). When model fit is insufficient, exploratory factor analysis (EFA) will be employed to examine potential multidimensionality. McDonald’s Omega will be computed to assess scale reliability. To evaluate the latent distribution of each construct, semiparametric and non-parametric factor models will be estimated and compared to the standard CFA model using AIC and BIC. To test discriminant validity, pairwise EFAs and pairwise CFAs will be performed to determine whether subscales are empirically distinguishable. Finally, the latent structure of the full internalizing scale will be examined using Bayesian CFA and higher-order EFA, allowing identification of overarching factors consistent with the HiTOP framework.

The evaluation metrics described in Section III will be applied and reported throughout all development phases. For model training and evaluation, the real-world validated dataset will be divided using an 80:20 split for training and testing sets, respectively. To ensure comprehensive model analysis, we plan to conduct two additional evaluations. First, an ablation study will examine the individual contribution of each proposed component to overall performance. Second, hyperparameter analysis will investigate the impact of key architectural parameters, such as the number of embedding dimensions or the number of layers in the GNN and Transformer-based encoder.

In addition to regression analysis, we propose to conduct complementary experiments examining the contextual symptom representations learned by the model at the embedding level. These investigations will assess the representational capacity of the learned symptom embeddings in capturing and encoding contextual information beyond what is revealed through decoded risk level predictions. While the specific experimental protocols for evaluating embedding quality and contextual sensitivity remain under development, these analyses will provide deeper insights into the model’s internal representation learning mechanisms and validate whether the learned embeddings effectively encode the complex interdependencies inherent in psychopathological symptom networks, especially for the development of the large language model in Phase 3.

### C. Phase 3 - Deployment

Following the completion of model training, we propose to integrate the validated framework into a clinical application designed to assist healthcare practitioners in establishing individualized psychopathology profiles for patients. The primary utility of this diagnostic tool lies in its capacity to reduce the assessment burden by enabling clinicians to input a limited set of critical symptom indicators while allowing the model to predict unmeasured symptom dimensions. Furthermore, the system leverages learned comorbidity patterns to identify and correct potential response inconsistencies, thereby enhancing diagnostic accuracy.

To advance this methodology further, the research agenda includes the development of a large language model capable of generating adaptive survey questions based on patients’ current responses. This dynamic questioning approach will be informed by the emerging symptom network structure and the centrality of symptoms within that network, potentially optimizing the diagnostic workflow and improving clinical efficiency.

The clinical application will undergo deployment across participating medical institutions for real-world validation. During the beta implementation phase, a comparative evaluation will be conducted to assess diagnostic process performance both with and without the assistance of the proposed tool. This evaluation framework will provide empirical evidence regarding the clinical utility and effectiveness of the AI-assisted diagnostic approach compared to traditional assessment methodologies.

## V. Strengths and Challenges

The development of an AI-enhanced individualized prediction tool for psychopathology profiling in Vietnam presents both significant opportunities for advancing mental health care and substantial implementation challenges that need careful considerations. This section examines the key advantages and potential obstacles associated with this research initiative.

### A. Strengths

Our study offers several distinctive advantages that position it as a pioneering contribution to the field of mental health digitalization and cross-cultural psychopathology assessment.

This work represents the first attempt to develop an AI-enhanced tool for predicting psychopathological traits based on the combined principles of both dimensional (HiTOP) and network approaches to psychopathology. This integration leverages the hierarchical structure of HiTOP while incorporating the dynamic relationships captured by network models, creating a more comprehensive framework for understanding mental health conditions.

The research provides a practical tool designed to support clinicians in mental health detection, periodic evaluation, monitoring intervention effectiveness, and tracking remission progress. This clinical utility addresses real-world needs in healthcare settings where accurate and efficient assessment tools can significantly improve patient outcomes and treatment planning.

Furthermore, the project advocates for cross-cultural adaptation of data-driven and scientific frameworks of psychopathology, which incorporate a less stigmatized and more data-driven view on mental disorders. This approach promotes evidence-based understanding of mental health conditions while reducing cultural barriers to treatment and assessment.

### B. Challenges

Despite these strengths, the implementation of this research faces several significant obstacles that require careful consideration and strategic planning.

The primary challenge involves deploying a new framework of psychopathology in Vietnam, a country where understanding and expertise in psychology remain limited. The current mental health infrastructure and professional training programs may not be adequately prepared to support the implementation of advanced dimensional assessment approaches.

Additionally, there are inherent challenges related to an etic approach of using frameworks developed in Western, high-income contexts within an Eastern, low-and-middle-income environment. Cultural differences in symptom expression and conceptualizations of mental health may affect the validity and applicability of the HiTOP framework in Vietnamese populations, requiring extensive validation and potential modifications to ensure cultural appropriateness and clinical utility.

## Data Availability

All data produced in the present study are available upon reasonable request to the authors

## VI. Future Work

This research will be extended in several key directions to maximize its clinical impact and scientific contribution. First, we will adopt a longitudinal design to model changes in symptom interactions over time and to identify early warning signals predictive of clinical deterioration and symptom development, thereby enhancing the tool’s temporal sensitivity and clinical utility. Additionally, we plan to scale deployment across clinical settings in Vietnam by integrating the tool as a standardized component of initial intake interviews, supporting early screening and individualized case formulation. To further advance the methodology, we propose leveraging analyses of symptom network structure and centrality to inform a dynamic questioning strategy, wherein a large language model generates adaptive survey items in real time, tailoring questions based on patients’ responses to prioritize the most informative items. This approach has the potential to improve the efficiency of diagnostic workflows and optimize clinical processes. Finally, we will explore the cross-cultural validity and generalizability of the HiTOP framework and symptom network approaches, thereby contributing to the global discourse on dimensional models of psychopathology in low- and middle-income countries.

## Acknowledgement

We would like to extend our gratitude to VinUni-Illinois Smart Health Center (VISHC) for organizing VISHC Pre-PhD Summer School 2024, which provided the environment and opportunity for our team to meet and initiate this project. We also thank our advisors and all those who contributed to the establishment of the project.

## Notes

### Competing Interest Statement

The authors have declared no competing interest.

### Funding Statement

This study has not received any funding yet.

### Author Declarations

The study will use theoretical frameworks in psychopathology and exisiting evidence on its parameters and distribution to generate synthetic data to train and test AI model.

## References

[1] Q. D. Tran, T. Q. C. Vu, and N. Q. Phan, “Depression prevalence in vietnam during the covid-19 pandemic: A systematic review and meta-analysis,” Ethics, Medicine and Public Health, vol. 23, p. 100806, 2022. [Online]. Available: https://www.sciencedirect.com/science/article/pii/S235255252200055X

[2] H. B. Nguyen, T. H. M. Nguyen, T. H. N. Vo, T. C. N. Vo, D. N. Q. Nguyen, H.-T. Nguyen, T.-N. Tang, T.-H. Nguyen, V. T. Do, and Q. B. Truong, “Post-traumatic stress disorder, anxiety, depression and related factors among covid-19 patients during the fourth wave of the pandemic in vietnam,” International Health, vol. 15, no. 4, pp. 365–375, 06 2022. [Online]. Available: 10.1093/inthealth/ihac040

[3] D. B. Olawade, O. Z. Wada, A. Odetayo, A. C. David-Olawade Asaolu, and J. Eberhardt, “Enhancing mental health with artificial intelligence: Current trends and future prospects,” Journal of Medicine, Surgery, and Public Health, vol. 3, p. 100099, 2024. [Online]. Available: https://www.sciencedirect.com/science/article/pii/S2949916X24000525

[4] M. L. Wainberg, P. Scorza, J. M. Shultz, L. Helpman, J. J. Mootz, K. A. Johnson, Y. Neria, J.-M. E. Bradford, M. A. Oquendo, and M. R. Arbuckle, “Challenges and opportunities in global mental health: a research-to-practice perspective,” Current psychiatry reports, vol. 19, pp. 1–10, 2017.

[5] A. Maqsood, S. Gul, T. Zahra, N. Noureen, and A. Khattak, “From face-to-face to screen-to-screen: exploring the multifaceted dimensions of digital mental health care,” Frontiers in Psychiatry, vol. 15, p. 1413127, 2024. [Online]. Available: 10.3389/fpsyt.2024.1413127

[6] J. Torous, J. Linardon, S. B. Goldberg, S. Sun, I. Bell, J. Nicholas, Hassan, Y. Hua, A. Milton, and J. Firth, “The evolving field of digital mental health: current evidence and implementation issues for smartphone apps, generative artificial intelligence, and virtual reality,” World Psychiatry, vol. 24, no. 2, pp. 156–174, 2025. [Online]. Available: https://onlinelibrary.wiley.com/doi/abs/10.1002/wps.21299

[7] T. T. Do, L. Van Tran, T. A. Le, T. M. Thi Le, L.-A. H. Duong, T. H. Nguyen, D. T. Phan, T. Van Vo, and H. T. Thi Ha, “Stress prediction using machine-learning techniques on physiological signals,” in 2023 1st International Conference on Health Science and Technology (ICHST), 2023, pp. 1–7.

[8] T. A. H. Nguyen, Q.-D. Nguyen, P. Pham, and L. T. T. Nguyen, “Enhancing machine learning approaches for early detection of depression levels for vietnamese students,” in Intelligent Systems Design and Applications, A. Abraham, A. Bajaj, and T. Hanne, Eds. Cham: Springer Nature Switzerland, 2024, pp. 388–397.

[9] American Psychiatric Association, Diagnostic and statistical manual of mental disorders: DSM-5, 5th ed. Washington, DC: Autor, 2013.

[10] W. H. Organization, “International classification of diseases, eleventh revision (icd-11),” 2018. [Online]. Available: https://icd.who.int/

[11] R. Kotov, R. F. Krueger, D. Watson, T. M. Achenbach, R. R. Althoff, R. M. Bagby, T. A. Brown, W. T. Carpenter, A. Caspi, L. A. Clark et al., “The hierarchical taxonomy of psychopathology (hitop): A dimensional alternative to traditional nosologies.” Journal of abnormal psychology, vol. 126, no. 4, p. 454, 2017.

[12] N. R. Eaton, L. F. Bringmann, T. Elmer, E. I. Fried, M. K. Forbes, A. L. Greene, R. F. Krueger, R. Kotov, P. D. McGorry, C. Mei et al., “A review of approaches and models in psychopathology conceptualization research,” Nature Reviews Psychology, vol. 2, no. 10, pp. 622–636, 2023.

[13] E. I. Fried and R. M. Nesse, “Depression is not a consistent syndrome: An investigation of unique symptom patterns in the star*d study,” Journal of Affective Disorders, vol. 172, pp. 96–102, 2015. [Online]. Available: https://www.sciencedirect.com/science/article/pii/S0165032714006326

[14] R. F. Krueger and N. R. Eaton, “Personality traits and the classification of mental disorders: Toward a more complete integration in dsm–5 and an empirical model of psychopathology.” Personality Disorders: Theory, Research, and Treatment, vol. 1, no. 2, p. 97, 2010.

[15] D. Borsboom, A. O. J. Cramer, V. D. Schmittmann, S. Epskamp, and L. J. Waldorp, “The small world of psychopathology,” PLOS ONE, vol. 6, no. 11, pp. 1–11, 11 2011. [Online]. Available: 10.1371/journal.pone.0027407

[16] D. A. Regier, W. E. Narrow, D. E. Clarke, H. C. Kraemer, S. J. Kuramoto, E. A. Kuhl, and D. J. Kupfer, “Dsm-5 field trials in the united states and canada, part ii: test-retest reliability of selected categorical diagnoses,” American journal of psychiatry, vol. 170, no. 1, pp. 59–70, 2013.

[17] W. Kim, Y. S. Woo, J.-H. Chae, and W.-M. Bahk, “The diagnostic stability of dsm-iv diagnoses: An examination of major depressive disorder, bipolar i disorder, and schizophrenia in korean patients,” Clinical Psychopharmacology and Neuroscience, vol. 9, no. 3, p. 117, 2011.

[18] I. R. Galatzer-Levy and R. A. Bryant, “636,120 ways to have posttraumatic stress disorder,” Perspectives on Psychological Science, vol. 8, no. 6, pp. 651–662, 2013, pMID: 26173229. [Online]. Available: 10.1177/1745691613504115

[19] E. Feczko, O. Miranda-Dominguez, M. Marr, A. M. Graham, J. T. Nigg, and D. A. Fair, “The heterogeneity problem: approaches to identify psychiatric subtypes,” Trends in cognitive sciences, vol. 23, no. 7, pp. 584–601, 2019.

[20] A. F. Marquand, T. Wolfers, M. Mennes, J. Buitelaar, and C. F. Beckmann, “Beyond lumping and splitting: a review of computational approaches for stratifying psychiatric disorders,” Biological psychiatry: cognitive neuroscience and neuroimaging, vol. 1, no. 5, pp. 433–447, 2016.

[21] M. J. Kozak and B. N. Cuthbert, “The nimh research domain criteria initiative: background, issues, and pragmatics,” Psychophysiology, vol. 53, no. 3, pp. 286–297, 2016.

[22] S. S. Kety, D. Rosenthal, P. H. Wender, and F. Schulsinger, “Mental illness in the biological and adoptive families of adopted schizophrenics,” American Journal of Psychiatry, vol. 128, no. 3, pp. 302–306, 1971, pMID: 5570994. [Online]. Available: 10.1176/ajp.128.3.302

[23] T. D. Cannon, P. M. Thompson, T. G. M. van Erp, A. W. Toga, V.-P. Poutanen, M. Huttunen, J. Lonnqvist, C.-G. Standerskjold-Nordenstam, K. L. Narr, M. Khaledy, C. I. Zoumalan, R. Dail, and J. Kaprio, “Cortex mapping reveals regionally specific patterns of genetic and disease-specific gray-matter deficits in twins discordant for schizophrenia,” Proceedings of the National Academy of Sciences, vol. 99, no. 5, pp. 3228–3233, 2002. [Online]. Available: https://www.pnas.org/doi/abs/10.1073/pnas.052023499

[24] S. E. Hyman, “Can neuroscience be integrated into the dsm-v?” Nature Reviews Neuroscience, vol. 8, no. 9, pp. 725–732, 2007.

[25] A. N. Goldstein-Piekarski, T. M. Ball, Z. Samara, B. R. Staveland, A. S. Keller, S. L. Fleming, K. A. Grisanzio, B. Holt-Gosselin, P. Stetz, J. Ma et al., “Mapping neural circuit biotypes to symptoms and behavioral dimensions of depression and anxiety,” Biological psychiatry, vol. 91, no. 6, p. 561, 2021.

[26] L. M. Williams, “Precision psychiatry: a neural circuit taxonomy for depression and anxiety,” The Lancet Psychiatry, vol. 3, no. 5, pp. 472–480, 2016. [Online]. Available: https://www.sciencedirect.com/science/article/pii/S2215036615005799

[27] D. C. Cicero, C. J. Ruggero, C. E. Balling, A. R. Bottera, S. Cheli, L. Elkrief, K. T. Forbush, C. J. Hopwood, K. G. Jonas, D. Jutras-Aswad, R. Kotov, H. F. Levin-Aspenson, S. N. Mullins-Sweatt, S. Johnson-Munguia, W. E. Narrow, S. Negi, C. J. Patrick, C. Rodriguez-Seijas, S. Sheth, L. J. Simms, and M. L. Thomeczek, “State of the science: The hierarchical taxonomy of psychopathology (hitop),” Behavior Therapy, vol. 55, no. 6, pp. 1114–1129, 2024, special Issue: State of the Science in Behavior Therapy: Taking Stock and Looking Forward. [Online]. Available: https://www.sciencedirect.com/science/article/pii/S0005789424000637

[28] N. R. Eaton, R. F. Krueger, K. E. Markon, K. M. Keyes, A. E. Skodol, Wall, D. S. Hasin, and B. F. Grant, “The structure and predictive validity of the internalizing disorders.” Journal of abnormal psychology, vol. 122, no. 1, p. 86, 2013.

[29] R. Kotov, D. C. Cicero, C. C. Conway, C. G. DeYoung, A. Dombrovski, R. Eaton, M. B. First, M. K. Forbes, S. E. Hyman, K. G. Jonas, and et al., “The hierarchical taxonomy of psychopathology (hitop) in psychiatric practice and research,” Psychological Medicine, vol. 52, no. 9, p. 1666–1678, 2022.

[30] D. J. Robinaugh, R. H. A. Hoekstra, E. R. Toner, and D. Borsboom, “The network approach to psychopathology: a review of the literature 2008–2018 and an agenda for future research,” Psychological Medicine, vol. 50, no. 3, p. 353–366, 2020.

[31] R. J. McNally, “Can network analysis transform psychopathology?” Behaviour Research and Therapy, vol. 86, pp. 95–104, 2016, contributions from experimental psychopathology to the understanding and treatment of mental disorders. [Online]. Available: https://www.sciencedirect.com/science/article/pii/S0005796716301103

[32] E. I. Fried and A. O. J. Cramer, “Moving forward: Challenges and directions for psychopathological network theory and methodology,” Perspectives on Psychological Science, vol. 12, no. 6, pp. 999–1020, 2017, pMID: 28873325. [Online]. Available: 10.1177/1745691617705892

[33] S. Epskamp, M. Rhemtulla, and D. Borsboom, “Generalized network psychometrics: Combining network and latent variable models,” Psychometrika, vol. 82, no. 4, p. 904–927, 2017.

[34] D. Borsboom, M. K. Deserno, M. Rhemtulla, S. Epskamp, E. I. Fried, R. J. McNally, D. J. Robinaugh, M. Perugini, J. Dalege, G. Costantini et al., “Network analysis of multivariate data in psychological science,” Nature Reviews Methods Primers, vol. 1, no. 1, p. 58, 2021.

[35] M. K. Deserno, M. S. M. Sachisthal, S. Epskamp, and M. E. J. Raijmakers, “A magnifying glass for the study of coupled developmental changes: Combining psychological networks and latent growth models,” Jan 2021. [Online]. Available: osf.io/preprints/psyarxiv/ngfxqv1

[36] H. F. Golino and S. Epskamp, “Exploratory graph analysis: A new approach for estimating the number of dimensions in psychological research,” PLOS ONE, vol. 12, no. 6, pp. 1–26, 06 2017. [Online]. Available: 10.1371/journal.pone.0174035

[37] H. Golino, D. Shi, A. P. Christensen, L. E. Garrido, M. D. Nieto, R. Sadana, J. A. Thiyagarajan, and A. Martinez-Molina, “Investigating the performance of exploratory graph analysis and traditional techniques to identify the number of latent factors: A simulation and tutorial.” Psychological methods, vol. 25, no. 3, p. 292, 2020.

[38] A. Vaswani, N. Shazeer, N. Parmar, J. Uszkoreit, L. Jones, A. N. Gomez, L. u. Kaiser, and I. Polosukhin, “Attention is all you need,” in Advances in Neural Information Processing Systems, I. Guyon, U. V. Luxburg, S. Bengio, H. Wallach, R. Fergus, S. Vishwanathan, and R. Garnett, Eds., vol. 30. Curran Associates, Inc., 2017. [Online]. Available: https://proceedings.neurips.cc/paperfiles/paper/2017/file/3f5ee243547dee91fbd053c1c4a845aa-Paper.pdf

[39] J. Shen, Z. Shen, C. Xiong, C. Wang, K. Wang, and J. Han, “Taxoexpan: Self-supervised taxonomy expansion with position-enhanced graph neural network,” in Proceedings of The Web Conference 2020, ser. WWW ‘20. New York, NY, USA: Association for Computing Machinery, 2020, p. 486–497. [Online]. Available: 10.1145/3366423.3380132

[40] A. van den Oord, Y. Li, and O. Vinyals, “Representation learning with contrastive predictive coding,” ArXiv, vol. abs/1807.03748, 2018. [Online]. Available: https://api.semanticscholar.org/CorpusID:49670925

[41] K. Sinha, R. Jia, D. Hupkes, J. Pineau, A. Williams, and D. Kiela, “Masked language modeling and the distributional hypothesis: Order word matters pre-training for little,” in Proceedings of the 2021 Conference on Empirical Methods in Natural Language Processing, M.-F. Moens, X. Huang, L. Specia, and S. W.-t. Yih, Eds. Online and Punta Cana, Dominican Republic: Association for Computational Linguistics, Nov. 2021, pp. 2888–2913. [Online]. Available: https://aclanthology.org/2021.emnlp-main.230/

[42] D. Watson, M. K. Forbes, H. F. Levin-Aspenson, C. J. Ruggero, Y. Kotelnikova, S. Khoo, R. M. Bagby, M. Sunderland, P. Patalay, and R. Kotov, “The development of preliminary hitop internalizing spectrum scales,” Assessment, vol. 29, no. 1, pp. 17–33, 2022, pMID: 33794667. [Online]. Available: 10.1177/10731911211003976

[43] P. Cruchinho, M. D. López-Franco, M. L. Capelas, S. Almeida, P. M. Bennett, M. Miranda da Silva, G. Teixeira, E. Nunes, P. Lucas, and F. Gaspar, “Translation, cross-cultural adaptation, and validation of measurement instruments: a practical guideline for novice researchers,” Journal of Multidisciplinary Healthcare, pp. 2701–2728, 2024.

[44] W. H. Organization, 2022. [Online]. Available: https://www.unicef.org/vietnam/media/9831/file/Study%20on%20school-related%20factors%20impacting%20mental%20health%20and%20well-being%20of%20adolescents%20in%20Viet%20Nam.pdf

[45] W. Falcon and The PyTorch Lightning team, “PyTorch Lightning,” Mar. 2019. [Online]. Available: https://github.com/Lightning-AI/lightning

[46] L. Biewald, “Experiment tracking with weights and biases,” 2020, software available from wandb.com. [Online]. Available: https://www.wandb.com/

[47] J. Zimmermann, L. P. Wendt, H. Edelhoff, E. Wierzba, L. Fleck, D. C. Cicero, and U. Reininghaus, “Development and initial evaluation of the german version of the hierarchical taxonomy of psychopathology self-report (hitop-sr),” 2024.

